# nnDoseNet: Intuitive and Flexible Deep Learning Framework to Train and Evaluate Radiotherapy Dose Prediction Models

**DOI:** 10.1101/2025.03.21.25324413

**Authors:** Ho-hsin Chang, Joseph Harms, Rex Alexander Cardan, John B. Fiveash, Richard A. Popple, Carlos E. Cardenas

## Abstract

**Background:** Radiotherapy (RT) dose optimization is often labor-intensive, requiring repeated manual adjustments to achieve clinically acceptable plans.

**Purpose:** In this work, we introduce nnDoseNet, a deep learning framework designed to automate and streamline RT dose prediction.

**Methods:** Building on the nnU-Net segmentation engine, nnDoseNet adapts this architecture for dose regression by incorporating specialized loss functions (including dose–volume histogram terms) and multi-channel input (CT, targets, organs-at-risk, and body mask). It also supports clinically relevant evaluation metrics (e.g., gamma analysis and D95).

**Results:** We evaluated nnDoseNet on the OpenKBP challenge dataset comprising 340 head-and-neck cancer cases (240 for training and 100 for testing). Multiple hyperparameters (U-Net depth, patch size, batch size, and loss function) were tested. The best-performing configuration achieved a dose score of 2.579 and a DVH score of 1.540 on the test set—competitive with top submissions in the original challenge. Additional validation on an institutional cohort of 80 prostate cancer patients (45 training, 35 testing) demonstrated good agreement with clinical dose distributions (mean-squared error 0.817) and improved target coverage compared to clinical plans.

**Conclusion:** By offering automated data preprocessing, systematic model training, and robust dose evaluation—all within a single framework—nnDoseNet reduces the complexity of building and testing dose prediction models. It accommodates diverse prescription doses, organ-at-risk definitions, and hardware configurations, making it a suitable benchmark for multi-institutional research. With its balance of simplicity, flexibility, and performance, nnDoseNet aims to accelerate the development, comparison, and clinical integration of advanced AI-driven dose prediction methods in radiotherapy.

## 1) Introduction

Radiation therapy (RT) is one of the most common treatments for cancer, with an estimated 60% of cancer patients receiving radiation at some point during the course of therapy^1^. RT treatment planning is a complex and subjective process required for all cancer patients receiving radiation treatments. It involves creating a 3D model of a patient’s anatomy, identifying tumors to treat and organs to avoid. Planning is an iterative optimization process where the radiation fields are designed to reduce doses to healthy tissues/toxicities; at its core, it is a central process for the safe and successful administration of radiotherapy. This treatment planning process is labor intensive, often taking several weeks between initiation of planning and the patient’s first treatment. More recently, novel techniques have been proposed to automate individual tasks within the planning process to increase efficiency and to make plans more consistent between planners.^2–5^

Knowledge-based planning (KBP) is one of RT dose prediction approach that derives new treatment plans from existing ones to facilitate automated planning, typically employing machine learning techniques. Shiraishi et al ^2^ trained an artificial neural network with dominant features of the planning such as number of fields, distance to planning target volumes (PTV), and beam angle. When convolution neural networks (CNN) were put into practice, researchers established a KBP model by using CNN based deep learning (DL) methods. The most outstanding and popular CNN based architecture is U-Net and its variants. In 2019, Nguyen et al^3^, developed a hierarchically densely connected U-Net trained with 120 head and neck cancer treatment plans which each contained 22 OARs and 1 to 5 PTVs ranging from 42.5 to 72 Gy. Babier et al ^4^ developed and trained a 3D generative adversarial network (GAN) with 130 oropharyngeal cancer treatment plans which each contained 8 OARs and dose was prescribed at 56, 63 and 70 Gy.

Advances in dose prediction architectures have been paralleled with progress in loss functions designed for this task. Mean squared error (MSE) and mean absolute error (MAE) are the most used loss functions for RT dose prediction. However, the lack of clinical evaluation metrics, such as DVH-metrics, applied during training could generate dose predictions with low loss values but fail to meet prescription and/or clinical goals. Some studies have proposed novel loss functions that incorporate MSE or MAE with DVH characteristics^4–7^ to address this limitation. Nguyen et al^5^ introduced a differentiable DVH-loss utilizing the sigmoid function. Koike et al. ^6^ developed a structure loss which normalized dose within a structure and measure its absolute difference of each voxel. Wang et al.^7^ proposed a value-based DVH loss (vDVH) that applies MAE to a sorted dose distribution and criteria-based (cDVH) DVH loss to calculate targets’ D99%[Gy], D5%[Gy], and D1%[Gy] and dose differences in D1cc [Gy] and Mean [Gy] on OARs.

In 2020, Babier et al. ^8,9^ organized the OpenKBP challenge, which provided participants with preprocessed datasets and clinical relevant evaluation code for deep learning-based KBP researches. Grand challenges, such as OpenKBP, provide a standardized process and evaluation template, ensuring consistency and comparability across different approaches. These challenges not only establish a benchmark for existing methodologies but also drive the development of novel models and techniques by inspiring participants to push the boundaries of innovation in the field^10–12^. A report summarizing the OpenKBP challenge^9^ showed that most of the challenge participants utilized a U-Net based approach. While many variants of the U-Net have been proposed, recent reports suggest that these variants often don’t improve performance. Isensee et al ^13^ introduced the nnU-Net, standing for “no new” U-Net, and demonstrated that a basic U-Net model proves to be sufficiently reliable for medical image segmentation, suggesting that the inclusion of specialized variant layers does not significantly enhance overall training performance. The nnU-Net has been widely investigated in research and clinical settings^14–19^, demonstrating that a well-designed tool that is flexible and easy to use can be widely accepted and can be applied to for a variety of segmentation tasks.

Despite advances in dose prediction research, significant barriers remain in widening applicability across different studies and institutions. These barriers arise from variations in radiotherapy planning, including differences in computing hardware, institutional protocols, treated sites, and delivery machines and indicate that a single trained model may not be general enough to address the diversity in clinical settings. Therefore, an intuitive framework that systematically train multiple DL models for different scenarios is important.

The OpenKBP challenge and nnU-Net exemplify how standardized, systematic frameworks can drive innovation in their respective research fields. Beyond serving as benchmarks, such frameworks provide structured workflows that allow researchers to focus on specific areas of interest without building projects from the ground up. Additionally, they offer a guided starting point for newcomers, reducing reliance on trial-and-error approaches and fostering more reproducible and efficient research. While nnU-Net is a segmentation framework and OpenKBP provides a template for deep learning-based dose prediction training, neither is designed for seamless integration with clinical datasets, which are typically stored in DICOM format. Users must handle key preprocessing steps, including formatting clinical data and converting predicted plans back into DICOM. Given the variability and inconsistencies in clinical datasets, this adds complexity to model development and deployment. Therefore, a comprehensive framework for radiotherapy dose prediction is needed—one that streamlines data preprocessing, model training, and integration with clinical datasets to improve workflow efficiency and reproducibility.

The primary aim of this study is to develop nnDoseNet, a generalized, flexible, and comprehensive framework for deep learning-based dose prediction. nnDoseNet is designed to accommodate a wide range of clinical scenarios, including various dose prescriptions, multiple target volumes, diverse beam geometries, and specific organ-at-risk (OAR) constraints. In terms of usability, it supports both clinical and well-structured datasets, operates on GPU-limited hardware, and is accessible to users with varying levels of expertise. To demonstrate its utility, we evaluate models trained with dose-prediction-specific loss functions across multiple disease sites and datasets.

## 2) Materials and Methods

### a. nnDoseNet

nnDoseNet builds on the approach of nnU-Net by automating the configuration process and adapting it specifically for radiotherapy dose prediction. In summary, we adapted the nnU-Net engine from segmentation tasks (i.e., classification) to regression tasks. Specific to radiotherapy dose prediction, we incorporate clinical dose evaluation features into model training using DVH-based loss functions. Additionally, we include clinical evaluation methods such as gamma analysis and DVH metrics calculation. Moreover, we standardize the RT structure extraction workflow, allowing users to utilize DICOM format datasets and integrate clinical goals and prescription information into the dataset for more clinically customized training.

nnDoseNet offers a flexible and intuitive framework for high-performance deep learning model training. Inspired by nnU-Net’s design principles, it enables users to customize key hyperparameters by modifying JSON configuration files—a human-readable, easily editable format that controls training and validation processes. This design ensures accessibility for users of all experience levels, allowing them to build, run, and fine-tune deep learning models using predefined commands and configuration files, without requiring direct code modifications.

The nnDoseNet workflow (Figure 1) consists of four main stages: (1) conversion, (2) preprocessing, (3) training, and (4) prediction. Each stage is driven by predefined commands and controlled through user-editable JSON configuration files (structure.json, dataset.json, and plan.json). These configuration files are automatically generated for each stage to ensure seamless progression through the workflow. The output of each stage serves as the input for the next, resulting in five structured operation folders: nnDose_dicom, raw, preprocess, results, and prediction. This structured, intuitive design enhances usability and generalizability across different clinical datasets and models.

**Figure 1.**
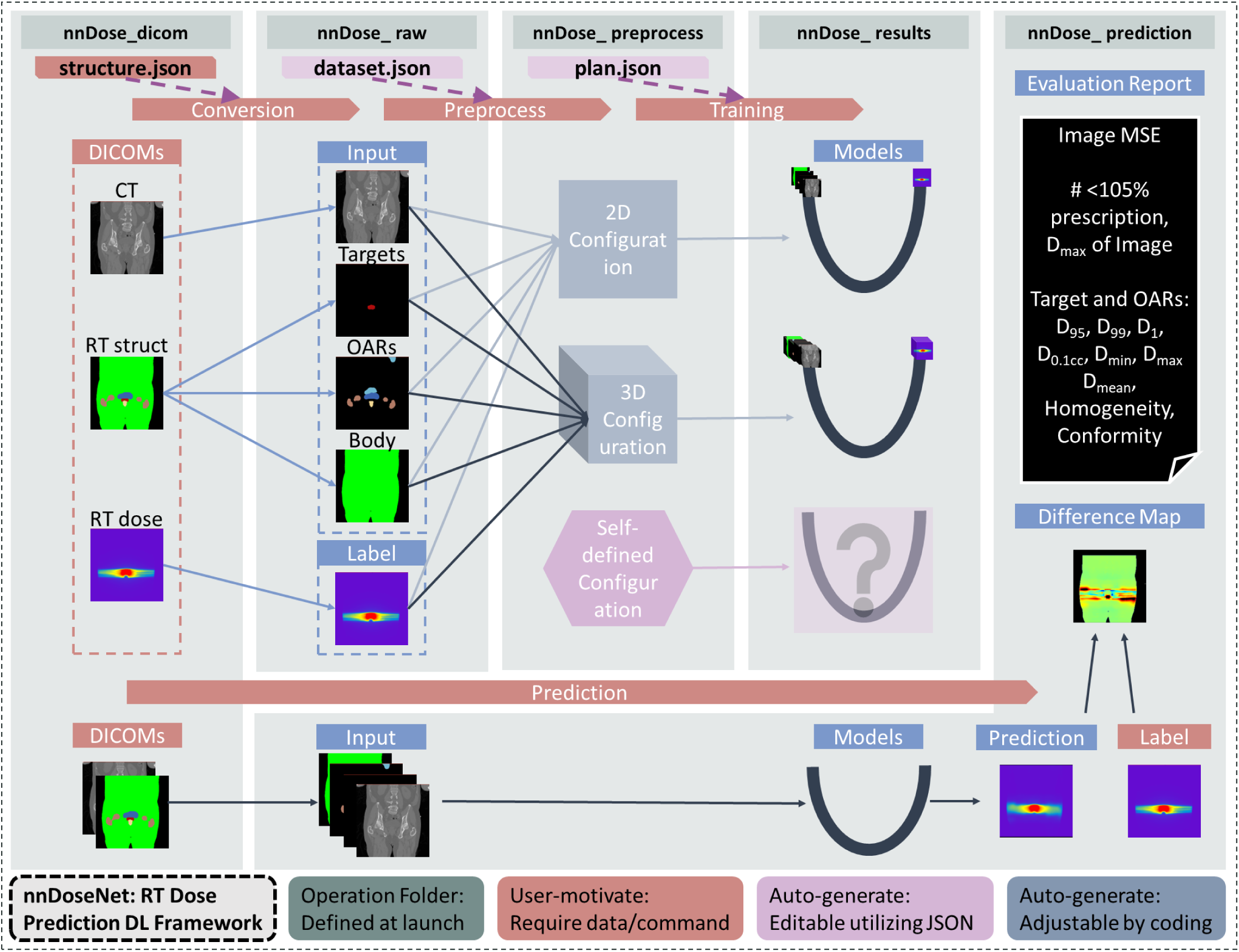
nnDoseNet workflow of 4 stages of operation and 5 operation folders (green). Training dataset (Tr) is involved in the first 3 operations while testing dataset (Ts) is only involved in the initial and final stages of operation. The red blocks mean the data need to be provided by user and the process need to be initial by user using predefined command(e.g. nnDoseNet require you to make command of nnDose_train to start training). The purple blocks means nnDoseNet would auto-generate the file or configuration, but the user has a lot of to modify the correspond json fil to alter the process (e.g. If you want to train a 3D model with smaller epochs than 1000, you would need to change the value of epoch_num in plan.json, no need to code). The blue blocks means the user need to change the process by coding(e.g. If the user want to use other architecture than U-Net, they would need to change the script in nnDoseTrainer).

#### a.a. DICOM operations

To support both clinical and research applications, nnDoseNet includes a conversion pipeline that transforms clinical DICOM data into NIfTI (Neuroimaging Informatics Technology Initiative) format, encapsulating the CT image, dose map, target structures, OARs, and body mask. The structure.json file provides two key functions:

1. Standardizing structure extraction from DICOM RTSTRUCT files by mapping structures to candidate ROI names to accommodate variations in clinical nomenclature.
2. Assigning structure-specific metadata, including integer labels, prescription values, and classification as either a target or an OAR.

This conversion occurs in two steps:

1. Extracting and converting the CT image, dose map, target structures, and OARs into individual NIfTI binary masks.
2. Assigning labels to each structure and combining them into consolidated 3D arrays for targets and OARs, stored as single NIfTI files.

This two-step approach provides flexibility for scenarios where structures are already available in NIfTI format or require updates to OAR labels or prescription doses based on evolving guidelines. Ultimately, nnDoseNet accommodates both DICOM and NIfTI datasets, ensuring efficient extraction and standardization of essential information for dose prediction..

#### a.b. Formatted dataset, preprocessing and plan

For preprocessing, nnDoseNet follows the nnU-Net workflow, with the formatted dataset from the DICOM conversion stage serving as input. The dataset.json file—automatically generated during DICOM processing—stores dataset-specific attributes such as image size, spacing, and origin, and defines modality-specific normalization methods. Following nnU-Net, CT intensities are clipped between the 5th and 95th percentiles, while target, OAR, and body mask values remain unaltered.

The workflow follows nnU-Net’s “pipeline fingerprints” approach, classifying parameters as fixed, rule-based, or empirical. This consists of two steps:

1. Analysis: Extracts dataset characteristics (spacing, size, intensity distribution) and generates a dataset fingerprint stored in dataset.json.
2. Preprocessing: Uses the dataset fingerprint to preprocess input data and document model architecture configurations in plan.json.

While most rule-based parameters from nnU-Net are retained, nnDoseNet modifies the resampling interpolation method for targets, OARs, body masks, and dose maps. Additionally, plan.json allows customization of key parameters such as epoch number and loss functions, enabling users to select from predefined architectures (2D, 3D, or 3D low-resolution U-Net) or define custom configurations. A detailed parameter table is provided in Appendix A (Table S1).

### a.c. Training parameters

#### a.c.1 Model architecture

Models are trained using default nnU-Net architectures (2D and 3D U-Nets with deep supervision). nnDoseNet follows nnU-Net’s training strategy, utilizing ‘Poly’ learning rate policy, Stochastic Gradient Descent (SGD) with weight decay, and deep supervision. For data augmentation, nnDoseNet applies different interpolation methods to mask-based channels (targets, OARs, and body masks) and image data (CT). Image-based augmentations, such as noise addition, are applied only to CT images.

#### a.c.2 DVH-Loss functions

To incorporate dose-volume histogram (DVH) constraints, nnDoseNet implements two specialized loss functions. vDVH extracts and sorts dose values within individual structures to compute structure-specific MAE, mimicking the DVH’s emphasis on high-dose regions rather than spatial dose distribution. cDVH calculates dose differences at D99, D95, and D1 for targets and Dmean, D0.1cc for OARs. The mathematical formulations for both loss functions are provided in Appendix B. By default, nnDoseNet applies vDVH and cDVH to target and OAR volumes while using MSE within the body mask. A total of 10 predefined loss modes are available for selection, as detailed in Appendix C, Table S2.

### a.d nnDoseNet inference pipeline

Similarly to nnU-Net, nnDoseNet provides an inference pipeline for trained models. Our implementation of the nnDoseNet inference pipeline includes the DICOM to NIfTI conversion, data preparation, dose prediction, dose evaluation (if reference dose files are available) and conversion from NIfTI to DICOM RTDOSE format.

### b. Datasets

#### b.a. OpenKBP dataset

The OpenKBP challenge provides 340 (240 training and 100 testing) head-and-neck cancer patient datasets, each contains a preprocessed CT image, PTV masks, OAR masks, body mask, and ground truth dose array. Each case has 1 to 3 targets and targets are categorized as low-, medium-, and high-risk with prescription doses of 56, 63 and 70 Gy, respectively. OARs include contours of brainstem, spinal cord, right and left parotid glands, larynx, esophagus, and mandible. However, not every case includes a full set of OAR contours.

#### b.b. Institutional Dataset

To further demonstrate the feasibility of our framework, we also evaluate its utility on clinical data from 80 (45 training and 35 testing) prostate cancer patients previously treated to a prescription of 70 Gy in 28 fractions using 2 or 3 full arcs VMAT at our institution. DICOM data was exported from our institution’s treatment planning system (TPS) and converted to NIfTI. Each patient’s data includes CT, structure set, and corresponding dose distribution files.

Structure sets included contours for targets, bladder, rectum, right and left femoral heads, bowel bag, and body.

#### b.c. Contours input channels

To incorporate prescription information during training, we assigned targets their respective prescription doses and labeled OARs with structure-specific integer values. As a result, individual voxels within target and OAR masks were assigned representative values for OpenKBP and institutional datasets, respectively. For example, all voxels containing PTV_High (Rx: 70 Gy) were set to have a value of 70 for the array used as the targets input channel. After labeling the contours, we combined all the OARs into one channel and the targets into another. When target structures overlap (i.e. PTV_High and PTV_Low) priority is given to higher-risk volumes, meaning that voxel values shared between volumes are assigned to the higher risk target. Similarly, when OARs overlap, overlapping voxels are assigned by highest-ranking order, which is defined in the structure.json file. This resulted in a total of four input channels: CT, targets, OARs, and the body mask.

### c. Experiments

To maximize the potential of nnDoseNet, we fine-tuned its default parameters, tested its adaptability, and validated findings from previous OpenKBP challenge related studies. We conducted several experiments on training loss, epoch number, patch size, hardware settings, and batch size using the OpenKBP dataset. Furthermore, we evaluated its performance on our institutional dataset using configurations of 3D full-resolution.

### d. Evaluation of metrics and statistical analysis

#### d.a OpenKBP software evaluation

We evaluate our results using the evaluation algorithm provided by OpenKBP which provides a dose score focusing on full dose map and a DVH score focusing on dose of targets and OARs. For a detailed explanation of these scores, we refer the reader to the manuscript by Babier et al^8^. In summary, the dose score is based on voxel-based MAE excluding the area outside the dose mask. The DVH score is a combination of DVH-based metrics: for OARs the mean dose and D_0.1cc_ (highest dose to 0.1 cc of the volume) within OAR mask are evaluated, and targets are evaluated using D_99_, D_5_, and D_1_. All comparisons are performed between the challenge-provided “ground-truth” and submitted/predicted dose distributions. For these comparisons, we utilize the OpenKBP split of 100 testing cases (not included in our training) and evaluate these using the OpenKBP-provided algorithm.

#### d.b nnDoseNet pipeline evaluation

For the institutional dataset, we use the built-in nnDoseNet evaluation pipeline which calculates MSE between the predicted and ground-truth dose and provides several clinically-relevant metrics such as D_min_, D_max_, D_mean_ and D_0.1cc_ for each structure and additional metrics for targets including D_1_, D_95_, D_99_, homogeneity and conformity.

For additional evaluation, nnDoseNet can generate a difference map that subtract voxel- to voxel intensity of prediction from ground-truth dose distributions. When DICOM data is available, nnDoseNet can write the prediction into DICOM format and calculate gamma analysis using a default of 3%/2mm threshold with 20 percent dose cutoff.

In this experiment, we have a held-out set of 35 cases (out of the original 80 cases) which are evaluated using the nnDoseNet evaluation pipeline. Here, we evaluate only the 3D U-Net model.

## 3) Results

We conducted a series of experiments on the OpenKBP dataset to fine-tune framework parameters and evaluated nnDoseNet on multiple disease sites using a clinical dataset to mimic real-world scenarios. The following experiments are performed using 2D and 3D U-Net architectures.

### i. Training loss

We defined 10 distinct loss mode combinations, integrating MSE or MAE with varying weights for vDVH and cDVH. Results from the OpenKBP test dataset (Table S2, Appendix C) indicate that (1) MSE-based loss functions outperform MAE-based loss functions, and (2) incorporating cDVH in the loss function leads to dose predictions that more closely align with ground-truth doses. Based on these findings, we focus the following experiments on loss functions that include cDVH, evaluating different weightings of vDVH and MSE.

### ii. Patch and batch sizes

We tested different patch and batch size combinations not just to find the best setting but also to mimic the scenario of training on GPUs with a range of VRAM. For our desktop scenario, we trained the models on Titan RTX with 24 GB VRAM available; for HPC applications we trained on A100 with 40 GB VRAM available. The experiment (Table S3 in Appendix C) shows that a patch size of [96,96, 64] outperformed other patch sizes, which is consistent with previous reports^11^. Increasing the batch size from 3 to 9 resulted in negligible performance improvement. Considering the efficiency gains, a batch size of 3 is ideal across all devices, whether using a desktop or HPC.

### iii. U-Net architecture – dimensions and depths

When comparing different model dimensions and depths (Table S4 in Appendix C), the 3D architecture outperformed 2D in terms of both dose and DVH scores. As for different depths in 3D U-Net the dose and DVH have small difference between depth of 4 to 5 while depth 5 has the best performance.

### iv. Optimal training parameters from previous experiments

Based on the above results (Table S2-4 in Appendix C), the best performing model used 3D-UNet with depth of 5 trained in loss mode 2 (0.5MSE+0.5cDVH) with patch size of [96,96,64] and batch size of 3. For a more rigorous evaluation, we also tested a depth of 6, loss mode 4 (0.6 MSE + 0.2 vDVH + 0.2 cDVH), and a batch size of 9, as these configurations yielded similar scores in previous experiments. The results, summarized in Table 1, indicate minimal differences in dose and DVH scores between these models.

**Table 1.** Dose score, DVH score and their combination score evaluated with OpenKBP evaluation algorithm comparing different parameters of model (depth, batch size and loss mode).

| Desktop |  |  |  |  |  | HPC |  |  |  |  |  |
| --- | --- | --- | --- | --- | --- | --- | --- | --- | --- | --- | --- |
| Depth | Batch Size | Loss | Dose Score | DVH Score | Dose +DVH | Depth | Batch Size | Loss | Dose Score | DVH Score | Dose +DVH |
| 5 | 3 | 2 | 2.602 | 1.528 | 4.129 | 5 | 9 | 2 | 2.618 | 1.558 | 4.177 |
| 6 | 3 | 2 | 2.584 | 1.538 | 4.122 | 6 | 9 | 2 | 2.592 | 1.535 | 4.127 |
| 5 | 3 | 4 | 2.605 | 1.537 | 4.142 | 5 | 9 | 4 | 2.619 | 1.537 | 4.156 |
| 6 | 3 | 4 | 2.579 | 1.540 | 4.119 | 6 | 9 | 4 | 2.590 | 1.552 | 4.142 |

**Table 2.** Summary of all experiments in experiments on OpenKBP dataset. In addition, we also include some of the default evaluation metrices of nnDoseNet, the number of cases have D_95_ of PTV_high above or equal prescription (70 Gy) and the number of cases has maximum dose of full dose map less than 105% of prescription (73.5 Gy) among 100 test cases in OpenKBP dataset. In ground truth (GT), there are 39/100 cases got D_95_ of PTV- high above prescription dose (70 Gy) and 57/100 cases have maximum dose in full dose map less than 105% prescription dose (73.5 Gy).

| *Experiment* | *Patch* | *Depth* | *Batch* | *Loss* | *Dose Score* | *DVH Score* | *Dose +DVH* | *# $D_{95} \geq$ Prescription*<br>(GT: 39/100) | *# <105% Prescription*<br>(GT: 57/100) |
| --- | --- | --- | --- | --- | --- | --- | --- | --- | --- |
| Loss | 128, 128, 80 | 6 | 3 | 2 | 2.605 | 1.540 | 4.145 | 27 | 88 |
| Loss | 128, 128, 80 | 6 | 3 | 4 | 2.620 | 1.552 | 4.173 | 26 | 87 |
| Depth | 128, 128, 80 | 5 | 3 | 3 | 2.615 | 1.541 | 4.156 | 29 | 88 |
| Depth | 128, 128, 80 | 6 | 3 | 3 | 2.624 | 1.553 | 4.177 | 29 | 86 |
| Patch, Batch | 96, 96, 64 | 6 | 3 | 3 | 2.586 | 1.541 | 4.127 | 31 | 90 |
| Patch, Batch | 96, 96, 64 | 6 | 9 | 3 | 2.589 | 1.536 | 4.125 | 29 | 86 |
| Winning | 96, 96, 64 | 5 | 3 | 2 | 2.602 | 1.528 | 4.129 | 28 | 89 |
| Winning | 96, 96, 64 | 6 | 3 | 4 | 2.579 | 1.540 | 4.119 | 31 | 91 |

### v. Summary and best model results

Table 3 summarizes superior models in each experiment stage. The best(lowest) dose score is achieved by models of same patch size [96,96,64], loss mode 4 and different batch size of 3. We follow OpenKBP challenge scoring and define our best model as the model achieved best(lowest) dose score. Figure 2 illustrates the comparison of dose distribution and DHV curves for a representative case. Figure 3 shows a boxplot of the nnDoseNet default dose evaluation metrics for all cases in the OpenKBP test set. Mean target and OAR doses are comparable between provided and predicted dose distributions.

**Figure 2.**
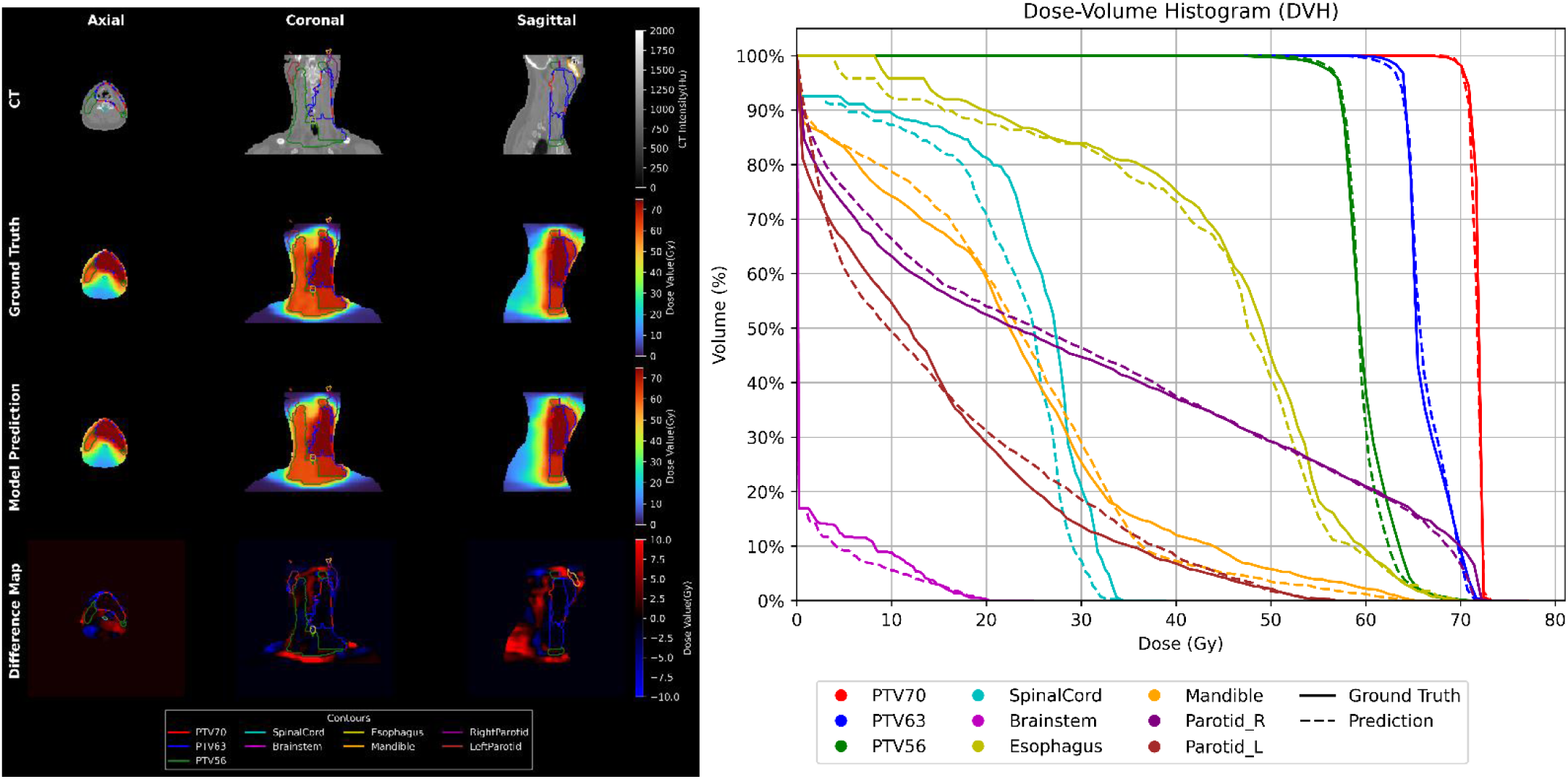
Left, difference map (ground truth subtracts prediction), dose map of ground truth and best dose score prediction (ensemble model of different batch size 3 with same parameter of depth of 6 trained in loss mode 4 and patch size [96,96,64]) of OpenKBP dataset. Up, DVH of ground truth (solid) and predicted (dash) dose distributions.

**Figure 3.**
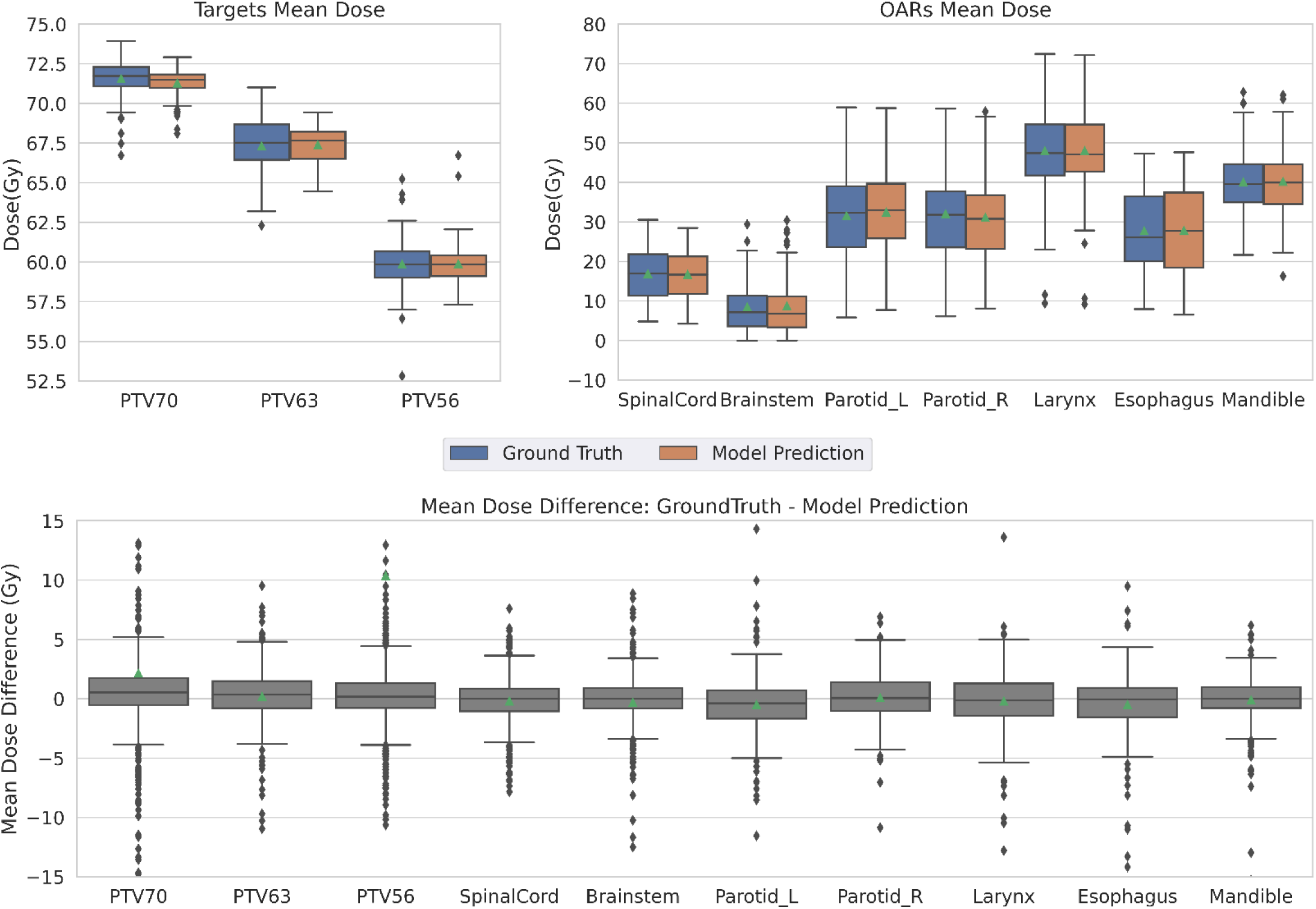
Boxplot of mean dose of PTVs(left) and OARs (right)). The ground truth (blue) and best dose score prediction model (orange) is ensemble model of different batch size 3 and 9 with same parameter of depth of 6 trained in loss mode 4 and patch size [96,96,64]. Other boxplots of nnDoseNet default evaluation metrics are in Figure S1 in Appendix D.

**Table 3.** Summary of top performance models in OpenKBP challenge and our model.

| Model | Dose Score | DVH Score | Dose+ DVH | Loss | Input channels | CT | targets | OAR | dose mask | Output (dose map) | Special |
| --- | --- | --- | --- | --- | --- | --- | --- | --- | --- | --- | --- |
| **Cascade 3D U-Net<sup>10</sup>** | 2.4294 | 1.4776 | 3.907 | MAE based (adapted to fit cascade model) | 9, 25 | Clipped -1024-1500 HU and normalized by 1000 Hu | Divided by highest prescription, and single channel | Binary masks, sperate channel | not mentioned | Divided by 70 Gy | deep supervision |
| **3D dense dilated U-Net <sup>11</sup>** | 2.5635 | 1.7045 | 4.268 | weighted-MSE(contour focused) | 12 | 0:3000 crop and rescaled to [0,1] | Binary masks, sperate channel | Binary masks, sperate channel | Binary masks, sperate channel | normalized to 70 Gy | [96,96,64] patch size |
| **CycleGAN(pix2pix)<sup>12</sup>** | 2.6497 | 1.5393 | 4.189 | MAE based (adapted to fit pix2pix model) | 11(from figure) | [0,4000] clip and rescale to [0,1] | not mentioned | not mentioned | not mentioned in input channel, but used for masking error | divided by 100 | pretrained model of pix2pix, masked MAE |
| **nnDoseNet (best dose score)** | 2.573 | 1.540 | 4.113 | MSE+vDVH +cDVH | 4 | 5th-95th percent ile clip | prescription dose, combined | clinical goal, combined | binary mask, sperate channel | none | deep supervision, median patch size, masked<br>loss. |

### vi. Institutional prostate dataset

The 3D model achieved a voxel-wise MSE of 0.817, and further analysis of clinical metrics revealed improved target coverage compared to the ground truth. Specifically, the model met D_95_ prescription dose coverage in 11 out of 35 cases, whereas the ground truth achieved this in only 3 out of 35 cases. Additionally, all predictions from the 3D model maintained a maximum dose below 105% of the prescription dose, compared to only 13 out of 35 cases in the ground truth. These findings suggest that the 3D model provides better target coverage while effectively minimizing extreme hotspots. As shown in Figure 4, the dose-volume histogram (DVH) curves for a representative case demonstrate improved target coverage and a steeper dose falloff compared to clinical plans, leading to lower D_max_ values in nnDoseNet predictions. However, as illustrated in Figure 5, dose predictions also resulted in higher-than-expected dose estimates for OARs, which may be a consequence of enhanced target coverage.

**Figure 4.**
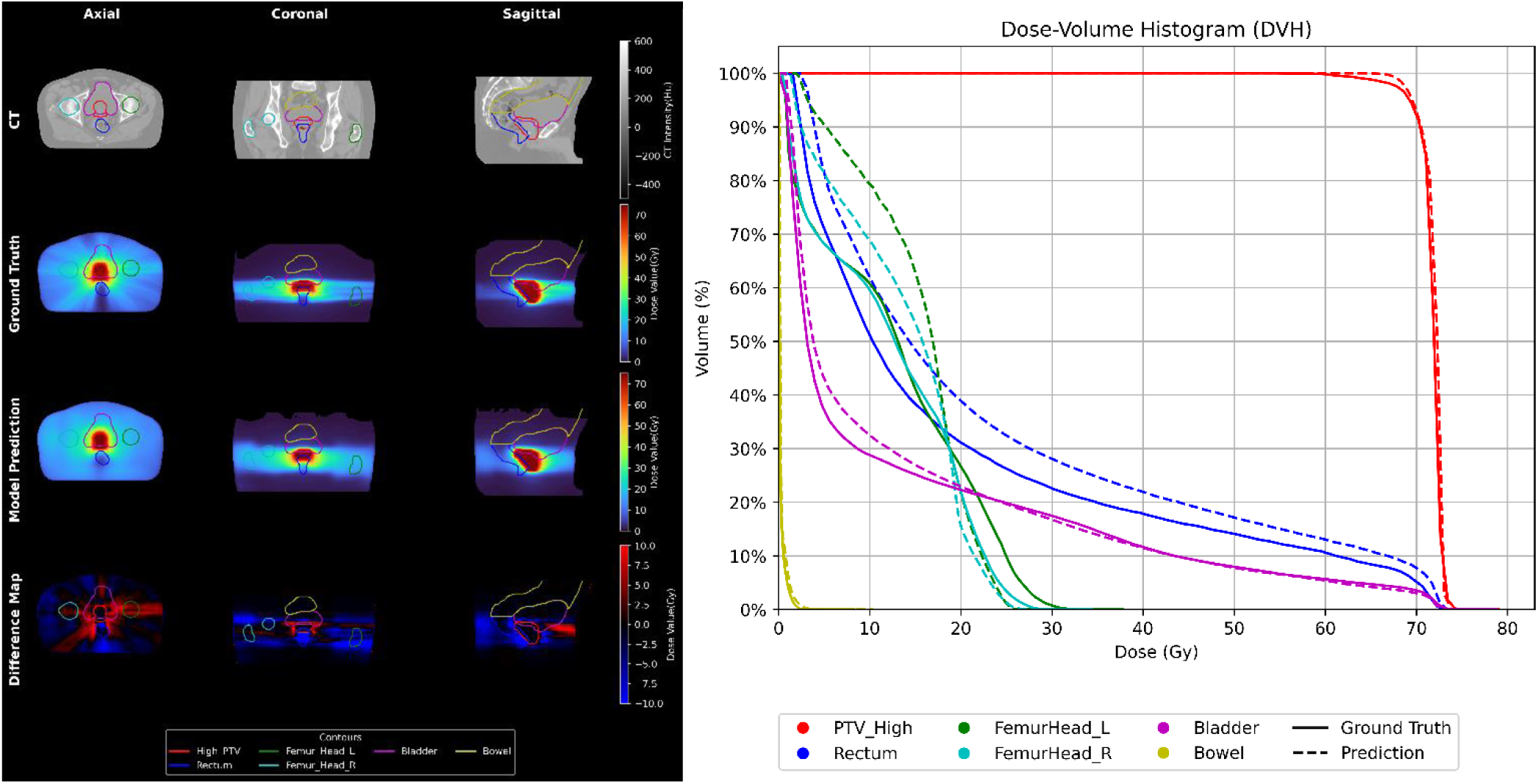
Left, difference map, dose map of ground truth and best prediction (3D full resolution) of institutional prostate dataset. Right, DVH of ground truth (solid) and predicted (dash) dose distributions.

**Figure 5.**
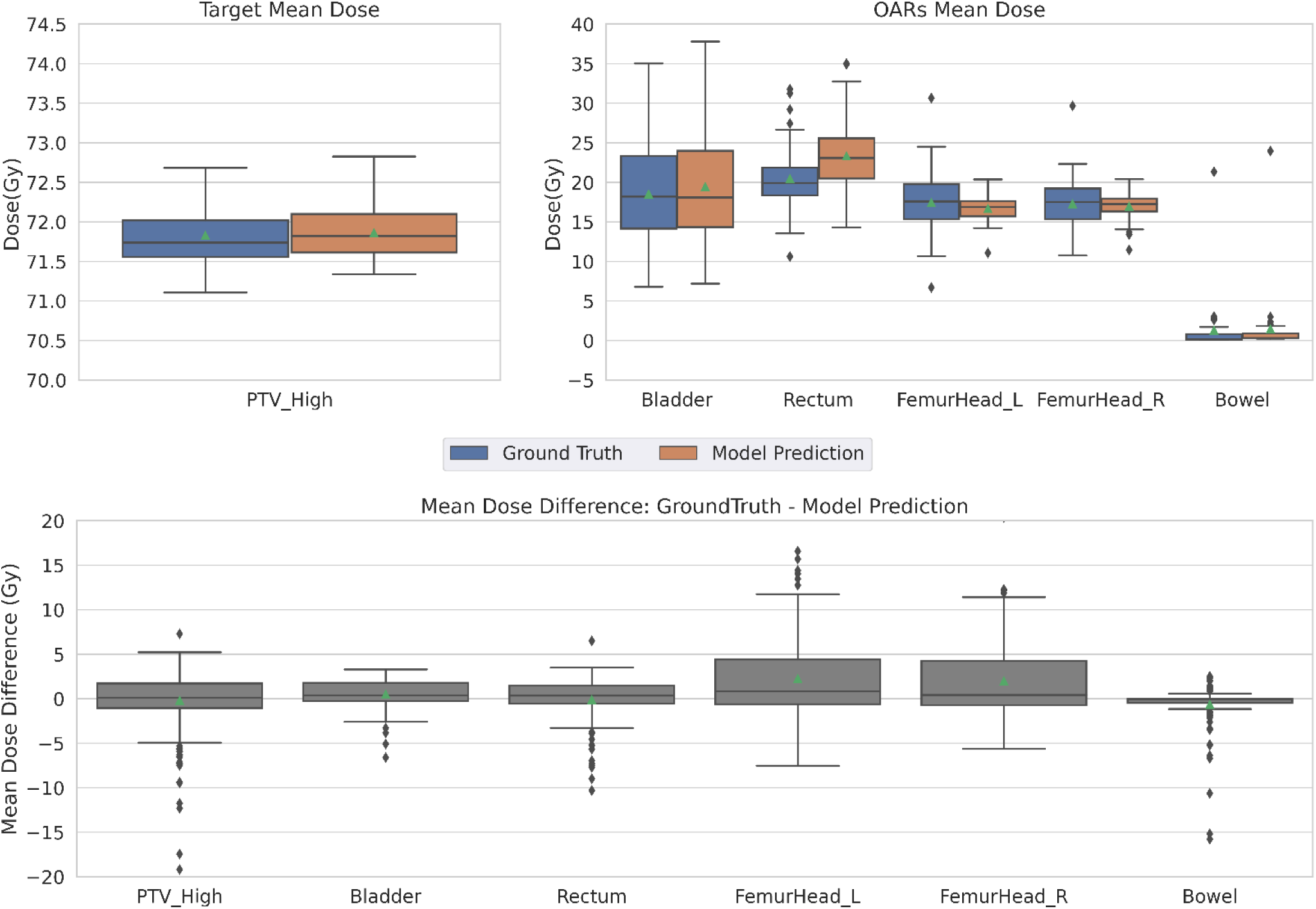
Boxplot of mean dose of PTV (left) and OARs (right)). The ground truth (blue) and best prediction model (orange) is the model trained with architecture of 3d U-Net. Other boxplots of nnDoseNet default evaluation metrics are Figure S2 in Appendix D.

## 4) Discussion

We introduced nnDoseNet, a deep learning-based framework designed to provide an off-the-shelf flexible, intuitive, and high-performance RT dose prediction tool. Installation of the nnDoseNet is straightforward, with options to install via <u>GitHub</u> or by downloading a Docker container. Despite its advanced capabilities, nnDoseNet remains user-friendly, requiring minimal coding experience. Most operations are streamlined into one or two commands, with further customization available through simple edits to the corresponding JSON files. Beyond its flexibility and ease of use, nnDoseNet has demonstrated good performance on both the OpenKBP challenge dataset and clinical data from our institution, highlighting its potential as a powerful tool for radiotherapy dose prediction research.

Upon comparing the results of our experiments, we found that the combination of loss functions incorporating MSE, vDVH, and cDVH yielded improved scores on the OpenKBP dataset. When assessing the impact of different U-Net depths, increasing a depth to 6 resolution steps indeed achieve better score than shallower models. However, the increase in score when moving from a depth of 5 to 6, is unlikely to be worth added computational time, prediction time, or model storage space.

In the experiment where we varied batch size and patch size, our aim was to simulate the hardware capabilities of typical workstations equipped with a single GPU and research HPCs (with many GPUs available) to identify a suitable patch size. As shown in Table S3 in Appendix C, a patch size of [96, 96, 64] outperformed most configurations, regardless of the batch size. This result aligns with the findings of Gronberg et al ^11^. However, nnDoseNet defaults to a median patch size because it provides a relatively good outcome and is versatile across different datasets. Factors such as spacing, image size, and the ratio of image background may influence the optimal patch size for training, thus warranting this median default.

Babier et al. ^4^ concluded that all OpenKBP participants used a variant model of either a U-Net or pix2pix, a GAN model with U-Net shape. The first place in both dose score and DVH score, the cascade 3DU-Net proposed by Liu et al^10^., is made with two 3D U-Nets with different initial feature. The first model gives a coarse prediction and passes the predictions as an input to the second model for fine-tuning. The second-best dose score was achieved by the 3D Dense Dilated U-Net developed by Gronberg et al.^6^ They employed a contour-focused loss function, the weighted-MSE loss, and specifically investigated the impact of patch size. The second place in DVH^12^ score is the one cycle learning pix2pix model with ResNet blocks, their experiment also applies masking to error in calculation in loss.

The top-performing nnDoseNet model achieved an OpenKBP dose score of 2.573, which would have won 3^rd^ place if the competition had still been open. Similarly, nnDoseNet’s best DVH-based submission would have achieved 2^nd^ place in the DVH score stream.

Our study has some limitations. First, the DICOM conversion pipeline is restricted to extracting dose maps from only one plan when multiple plans may be present, and it may miss structures if the user does not correctly set up the structure.json file. Additionally, the algorithm prioritizes the first structure name (most common name) in cases where multiple potential names are provided (e.g. PTV_High and PTV70 are both defined as high-risk PTVs within 1 structure set). Although a report is generated to indicate the extracted structure names, this limitation could impact the comprehensiveness of the data and must be carefully reviewed by users prior to moving on to the data preprocessing step. Another limitation of nnDoseNet is its lack of beam geometry information as an input, preventing the model from distinguishing between IMRT and VMAT cases. Incorporating beam geometry data into the training/testing process is a key area of future development, enabling the framework to handle mixed IMRT/VMAT datasets more effectively.

## 5) Conclusion

We developed nnDoseNet, a generalized and flexible framework for deep learning-based radiotherapy dose prediction. The framework integrates multiple dataset preparation methods (including customizable labeling and normalization), supports various model architectures (2D, 3D full-resolution, and ensembles), incorporates DVH-informed loss functions, and provides a comprehensive set of clinical evaluation metrics (e.g., gamma analysis, D95) within a single, unified system. We conducted a thorough evaluation of its hyperparameters using both public and institutional datasets, demonstrating its adaptability and strong performance across diverse clinical scenarios. These findings highlight nnDoseNet as a robust, scalable, and publicly available framework for advancing RT dose prediction research.

## Data Availability

This study utilized two data sources. The OpenKBP dataset, which is publicly available, can be accessed at GitHub. The second dataset, while not publicly available, can be made available upon reasonable request to the corresponding author, subject to appropriate institutional and ethical approvals.

https://github.com/ababier/open-kbp

