## appendix for "nnDoseNet: Intuitive and Flexible Deep Learning Framework to Train and Evaluate Radiotherapy Dose Prediction Models"

### Appendix A:

| Automated | Required input | Details (fixed, rule-based or empirical) configuration derived by distilling expert knowledge (more details in online methods) |
|---|---|---|
| Learning rate | – | Poly learning rate schedule (initial, 0.01) (default is 1000 epoch, which make the decay increment 1 e-5) |
| Architecture template | – | Encoder-decoder architecture with skip-connection. Other setting include instance normalization, leaky ReLU, deep supervision |
| Optimizer | – | SGD with Nesterov momentum (μ = 0.99) |
| Data augmentation | – | Rotations, scaling and mirroring on all channel. Gaussian noise, Gaussian blur, brightness, contrast, simulation of low resolution and gamma correction ONLY on non-mask(img, CT) channel. |
| Epoch number | | 1000(default), can be changed through plan.json |
| Loss function | – | masked-MAE or masked-MSE with different weight of vDVH and cDVH.(Loss modes definition in experiment training loss ) |
| Training procedure | – | epoch number× 250 minibatches, foreground oversampling |
| Inference procedure | – | Sliding window with half-patch size overlap, Gaussian patch center weighting |
| Intensity normalization | Modality, intensity distribution (auto-extract) | If CT, global dataset 5th-95th percentile clipping |
| Image resampling strategy | Distribution of spacings(auto-extract) | Third-order spline for CT and dose map |
| Mask resampling strategy | Distribution of spacings(auto-extract) | nearest neighbor for contours (PTV, OAR and body) contour |
| Image target spacing | Distribution of spacings(auto-extract) | If anisotropic, lowest resolution axis tenth percentile, other axes median. Otherwise, median spacing for each axis. (computed based on spacings found in training cases) |
| Network topology, patch size, batch size | Median resampled shape, target spacing, GPU memory limit(auto-extract, except GPU memory need manually define in command) | Initialize the patch size to median image shape and iteratively reduce it while adapting the network topology accordingly until the network can be trained with a batch size of at least 2 given GPU memory constraints. |
| Configuration of low-resolution 3D U-Net | Low-res target spacing or image shapes, GPU memory limit(auto-extract, except GPU memory need manually define in command) | Iteratively increase target spacing while reconfiguring patch size, network topology and batch size (as described above) until the configured patch size covers 25% of the median image shape. |
| Ensemble selection | Full set of training data and annotations (auto-extract) | From 2D U-Net, 3D U-Net, and all of their varient from adjustment to plan.json |

*Table 1 Table of fixed parameters (green), rule-based parameters (blue) and empirical parameters (purple). All of rule-based and empirical parameter can be easily changed through plan.json file or nnDoseNet command.*

#### Appendix B:

$n_x$: total numer of voxel in contour $x$

$N_x$: total numer of contour $x$

$contour$: include all PTV and all OAR

$Pr$: Percentile

$$vDVH_{contour} = \frac{\left|sorted\left[Mask_{contour} \times Dose_{gt}\right] - sorted\left[Mask_{contour} \times Dose_{predict}\right]\right|}{n_{contour}}$$

$$vDVH = Mean(vDVH_{ptv1} + vDVH_{ptv1} + \cdots + vDVH_{oar1} + vDVH_{oar2} + \cdots)$$

$$\begin{aligned} cDVH_{PTV} = \; & \left|Pr_{99}\left[Mask_{ptv} \times Dose_{gt}\right] - Pr_{99}\left[Mask_{ptv} \times Dose_{predict}\right]\right| \\ & + \left|Pr_{95}\left[Mask_{ptv} \times Dose_{gt}\right] - Pr_{95}\left[Mask_{ptv} \times Dose_{predict}\right]\right| \\ & + \left|Pr_{1}\left[Mask_{ptv} \times Dose_{gt}\right] - Pr_{1}\left[Mask_{ptv} \times Dose_{predict}\right]\right| \end{aligned}$$

$$\begin{aligned} cDVH_{OAR} = \; & \left|Mean\left[Mask_{oar} \times Dose_{gt}\right] - Mean\left[Mask_{oar} \times Dose_{predict}\right]\right| \\ & + \left|Pr_{1-\frac{1}{n_{oar}}}\left[Mask_{oar} \times Dose_{gt}\right] - Pr_{1-\frac{1}{n_{oar}}}\left[Mask_{oar} \times Dose_{predict}\right]\right| \end{aligned}$$

$$cDVH = \frac{cDVH_{ptv1} + cDVH_{ptv1} + \cdots + cDVH_{oar1} + cDVH_{oar2} + \cdots}{3 \times N_{ptv} + 2 \times N_{oar}}$$

$$MSE_{masked} = \frac{(Mask_{Body} \times Dose_{gt} - Mask_{Body} \times Dose_{predict})^2}{n_{Body}}$$

$$MAE_{masked} = \frac{\left|Mask_{Body} \times Dose_{gt} - Mask_{Body} \times Dose_{predict}\right|}{n_{Body}}$$

$*$ $x^2$ and $|x|$ in $MSE_{masked}$ and $MAE_{masked}$ are operate voxel by voxel

### Appendix C:

| 1. Loss Mode | MSE+DVH Combination | Dose Score | DVH Score | Dose +DVH | Loss Mode | MAE +DVH Combination | Dose Score | DVH Score | Dose +DVH |
|---|---|---|---|---|---|---|---|---|---|
| 0 | 1*MSE | 2.778 | 1.818 | 4.597 | 5 | 1*MAE | 2.622 | 1.541 | 4.162 |
| 1 | 0.5*MSE +0.5*vDVH | 2.627 | 1.556 | 4.183 | 6 | 0.5*MAE +0.5*vDVH | 2.629 | 1.555 | 4.185 |
| 2 | 0.5*MSE +0.5*cDVH | 2.605 | 1.540 | 4.145 | 7 | 0.5*MAE +0.5*cDVH | 2.621 | 1.547 | 4.168 |
| 3 | 0.4*MSE +0.3*vDVH+0.3*cDVH | 2.624 | 1.553 | 4.177 | 8 | 0.4*MAE +0.3*vDVH+0.3*cDVH | 2.634 | 1.563 | 4.197 |
| 4 | 0.6*MSE +0.2*vDVH+0.2*cDVH | 2.620 | 1.552 | 4.173 | 9 | 0.6*MAE +0.2*vDVH+0.2*cDVH | 2.628 | 1.551 | 4.180 |

*Table 2 Dose score, DVH score and their combination score evaluate with OpenKBP evaluation software comparing different loss mode which are set in nnDoseNet.*

| **Desktop** | | | | | **HPC** | | | | |
|---|---|---|---|---|---|---|---|---|---|
| Batch Size | Patch Size | Dose Score | DVH Score | Dose +DVH | Batch Size | Patch Size | Dose Score | DVH Score | Dose +DVH |
| 3 | 32, 32, 32 | 3.278 | 1.953 | 5.231 | 9 | 32, 32, 32 | 3.083 | 1.810 | 4.893 |
| 3 | 64, 64, 32 | 2.857 | 1.634 | 4.490 | 9 | 64, 64, 32 | 2.771 | 1.550 | 4.321 |
| 3 | 64, 64, 64 | 2.653 | 1.571 | 4.224 | 9 | 64, 64, 64 | 2.619 | 1.542 | 4.162 |
| 3 | 96, 96, 32 | 2.761 | 1.571 | 4.333 | 9 | 96, 96, 32 | 2.749 | 1.556 | 4.305 |
| 3 | 96, 96, 64 | 2.586 | 1.541 | 4.127 | 9 | 96, 96, 64 | 2.589 | 1.536 | 4.125 |
| 3 | 96, 96, 96 | 2.596 | 1.543 | 4.139 | 9 | 96, 96, 96 | 2.608 | 1.537 | 4.144 |
| 3 | 128, 128, 32 | 2.796 | 1.599 | 4.395 | 9 | 128, 128,32 | 2.831 | 1.590 | 4.422 |
| 3 | 128, 128, 64 | 2.611 | 1.561 | 4.172 | 9 | 128, 128, 64 | 2.676 | 1.587 | 4.263 |
| 3 | 128, 128, 80 | 2.615 | 1.56 | 4.175 | 9 | 128, 128, 80 | 2.661 | 1.568 | 4.229 |

*Table 3 Dose score, DVH score and their combination score evaluate with OpenKBP evaluation software comparing different patch size and different batch size for different usage scenarios*

| Dimension | Depth | Dose Score | DVH Score | Dose +DVH |
|---|---|---|---|---|
| 2D | 5 | 3.912 | 2.232 | 6.144 |
| 3D | 3 | 2.959 | 1.682 | 4.641 |
| 3D | 4 | 2.640 | 1.556 | 4.196 |
| 3D | 5 | 2.615 | 1.541 | 4.156 |
| 3D | 6 | 2.624 | 1.553 | 4.177 |

*Table 4 Dose score, DVH score and their combination score evaluate with OpenKBP evaluation software comparing different scores of 2D or 3D UNet model with different depth.*

### Appendix D:

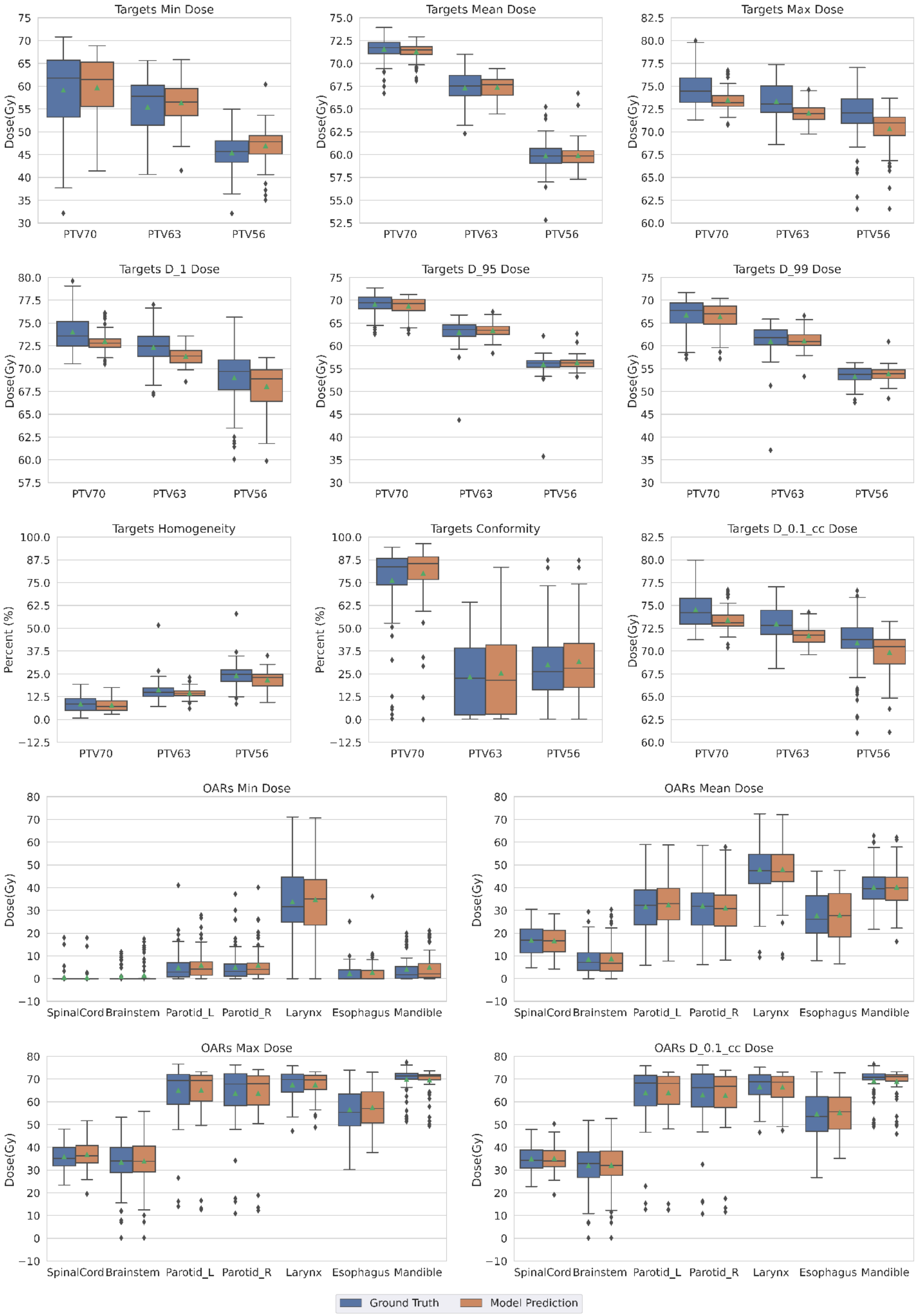


*Figure 1 Boxplot of all evaluation metrices nnDoseNet provides for targets and OARs. The ground truth (blue) and best prediction model (orange) is the ensemble model of different batch size 3 and 9 with same parameter of depth of 6 trained in loss mode 4 and patch size [96,96,64].*

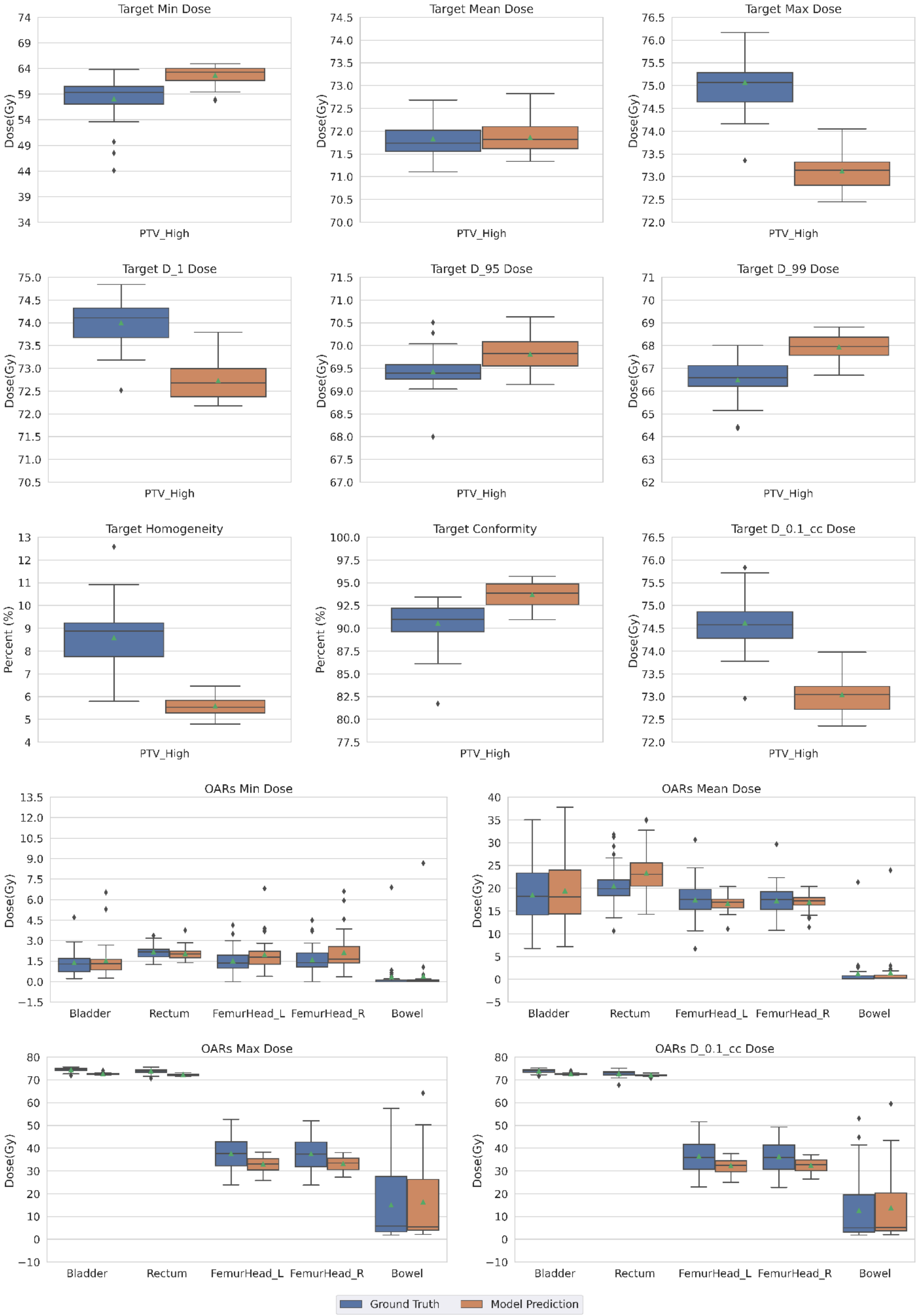


*Figure 2 Boxplot of all evaluation metrices nnDoseNet provides for targets and OARs. The ground truth (blue) and best prediction model (orange) is the model trained with architecture of 3d full resolution with default setting.*
